# Evaluating the effects of re-opening plans on dynamics of COVID-19 in São Paulo, Brazil^1^

**DOI:** 10.1101/2021.01.14.21249809

**Authors:** F.A. Rubio, T.N. Vilches

## Abstract

Coronavirus disease 2019 (COVID-19) was declared a pandemic by the World Health Organization in early March 2020. In Brazil, São Paulo is the most affected state, comprising about 20% of the country’s cases. With no vaccine available to date, distancing measures have been taken to reduce virus transmission. To reduce the pandemic’s effect on the economy, the government of São Paulo has proposed a plan consisting of five phases of the gradual re-opening of activities. In this context, we have developed a mathematical model to simulate the gradual re-opening plan on the transmission dynamics of COVID-19, in the city of São Paulo. The model shows that a precipitous reopening can cause a higher peak of the disease, which may compromise the local health system. Waiting for the reduction in the incidence of infected individuals for at least 15 days to phase transition is the most efficient strategy compared to the fixed-period scenario at each phase of the re-opening plan.

## 1. Introduction

In early 2020 the world started experiencing one of the most important and harmful pandemics of the last century, following the classification of World Health Organization (WHO) on March 11^*th*^ [1]. In July, the Coronavirus Disease - 2019 (COVID-19), illness whose pathological agent is the virus SARS-CoV-2, had already spread to all continents, caused more than 22,000,000 of cases, and more than 700,000 deaths [2]. In Brazil, the second most affected country until now, the spread started in São Paulo (SP) City, the most populous city in the country, where the first case was reported on February 26^*th*^ [3].

Given the current scenario of the pandemics, several institutions have been working on the vaccine against the COVID-19 [4, 5, 6]. However, it is estimated that a first candidate will be ready to be used on critical cases only in the early 2021 [6]. While the vaccine is not ready, and considering that there is no significant herd immunity in the population, the control measures have been, and must be, based on the social distancing and cases detection and isolation [7].

The government of São Paulo State designed strategies of control based on social distancing, closing all non-essential activities [8]. All non-essential business were kept close until end of May, when the government of SP proposed a re-opening plan composed of five phases: (1) the more restrictive phase, in which all non-essential business is closed; (2) malls, stores and services can operate with 20% of their capacity; (3) restaurants, pubs and hair saloons can open with 40% of their capacity; (4) Gyms are allowed to open with 60% of their capacity; and (5) All activities are allowed [9]. Also, moving from one phase to another, what was allowed in the previous phase remains in the next stage. On June 10^*st*^, the capital of the state, SP City had entered to the phase 2 of the re-opening plan and, June 29^*th*^, it started the phase 3 and has remained at that stage ever since [10].

We aim to investigate how the dynamics of COVID-19 may behavior due to re-opening stages in SP City. In order to do that, we fitted a classical SEIR model with a temporal transmission rate, *β*(*t*), to data provided by the government of SP State, using a genetic algorithm, and estimated the effective reproductive number, *R*_*t*_. After that, using the average of the last three weekly fitted values of *β*(*t*), we simulated different re-opening scenarios, and discussed the results of simulations.

## 2. Model formulation

The model proposed is subdivided into five epidemiological classes, composed by susceptible (*S*), exposed (*E*), infectious (*I*), recovered (*R*), and dead by COVID-19 (*D*), respectively. Since the epidemic occurs fast enough, demographic processes were not considered. Moreover, despite some few cases of immunity waning after infection, we assumed that the recovered population develops long-term immunity against COVID-19.

Susceptible people can be infected by COVID-19 at a disease transmission rate *β*, moving to the exposed compartment. In the compartment *E*, we consider that an exposed person cannot infect others, given that it is the incubation period of the virus. After an average period *α*^−1^, this person becomes an infectious individual, which can recover after a period *γ*^−1^. The disease-induced mortality is given by the term *θI*. In order to explore this effect, we consider *ρ* as the probability of dying due to COVID-19 during the infectious period, giving *θ* = *ρ*(1 − *ρ*)^−1^*γ* [11].

Therefore, we have a system of non-linear ordinary differential equations, given by

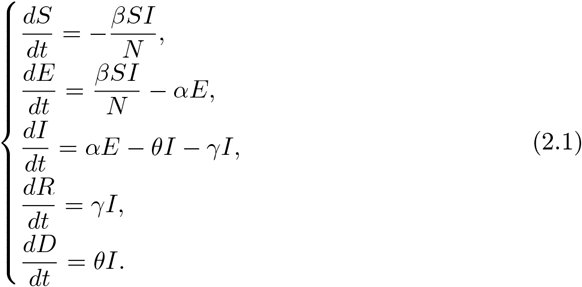

where *N* = *S* + *E* + *I* + *R*.

Table 1 summarises the model’s parameters, their description and values used in the simulations. Figure 1 shows the flow diagram of the dynamics.

**Tabela 1:**
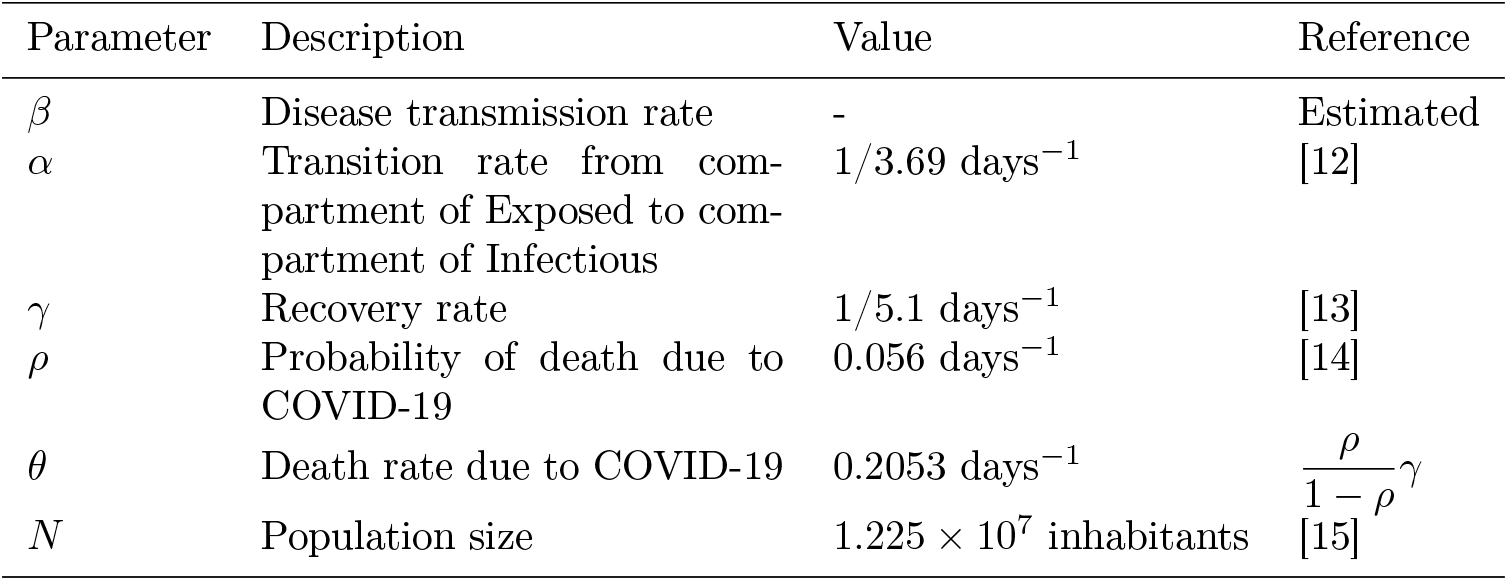
Parameters description of the model. All parameters have dimension day^−1^.

**Figura 1:**
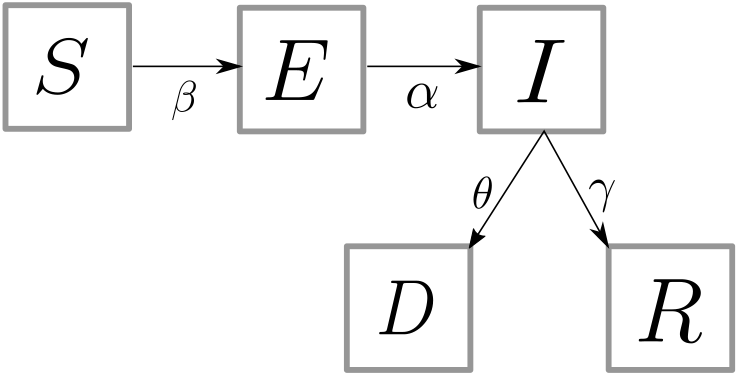
Diagram summarizing the interactions between variables of the model (2.1). The black arrows show the movement between the classes.

The last equation in the system (2.1) is not coupled with the other equations. Therefore, the system above can be rewritten to

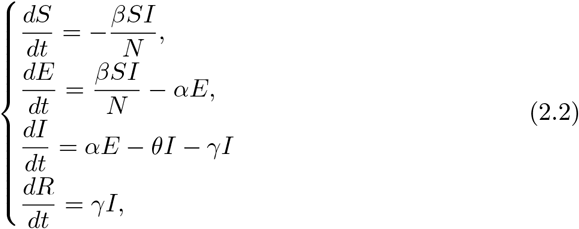

The number of deaths by COVID-19, *D*, can be computed as follows:

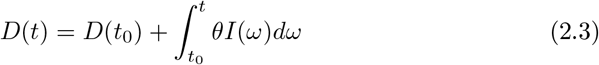

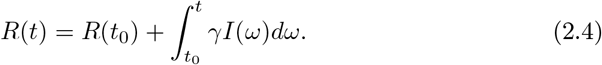

Thus, we can analyze a system with only three variables, which are *S, E*, and *I*.

## 3. Analysis of the model

We determine the disease-free equilibrium (DFE) point and the condition to occurs an epidemic peak, which gives us a threshold that we use to estimate *β*. Based on this parameter estimation, social-distancing measures were evaluated and discussed.

### 3.1. The threshold phenomenon

We denote an arbitrary equilibrium point as *P* = (*S, E, I, R*). DFE point is given by *P*^0^= (*N*, 0, 0, 0). Using the Next-Generation Matrix we assessed the condition of existence of the epidemic peak. For this, we considered the matrices of transmission and transition, 𝔽 and 𝕍, respectively, where

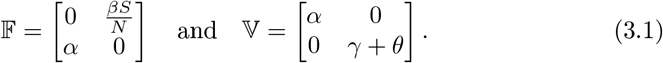

According to Van den Driessche et al. (2002), the next-generation matrix is

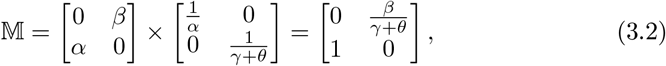

whose the characteristic polynomial is 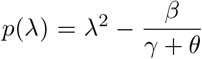. Using the methodology of Yang and Greenhalgh (2015), we obtain dimensionless parameter 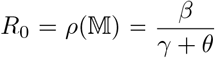, for which, if *R*_0_ < 1 then the diseases dies out, and if *R*_0_ > 1, there is an epidemic peak [16, 17, 18].

In epidemiology, the parameter *R*_0_ is known as the basic reproductive number and displays an important role. It can be interpreted as the number of secondary cases resulted from a primary case in an entirely susceptible population [11].

### 3.2. Parameter Estimation

Initially, we sought to estimate the parameter of transmission, *β*, using data from the government of São Paulo State [19]. The control measure, as it was told before, is implemented through social distancing and has a plan divided in phases. The social distancing index, calculating by the government using data from cellphone’s GPS, display several oscillations as shown in Figure 2a. The phases of the reopening plan show to reflect on the number of cases and, consequently, the number of deaths, as it is shown in figures 2b and 2c. We can see that, around 15 days before the begging of phase 3 in São Paulo City, the number of cases seems to display an increasing behavior, which may be followed by the increase in the number of deaths.

**Figura 2:**
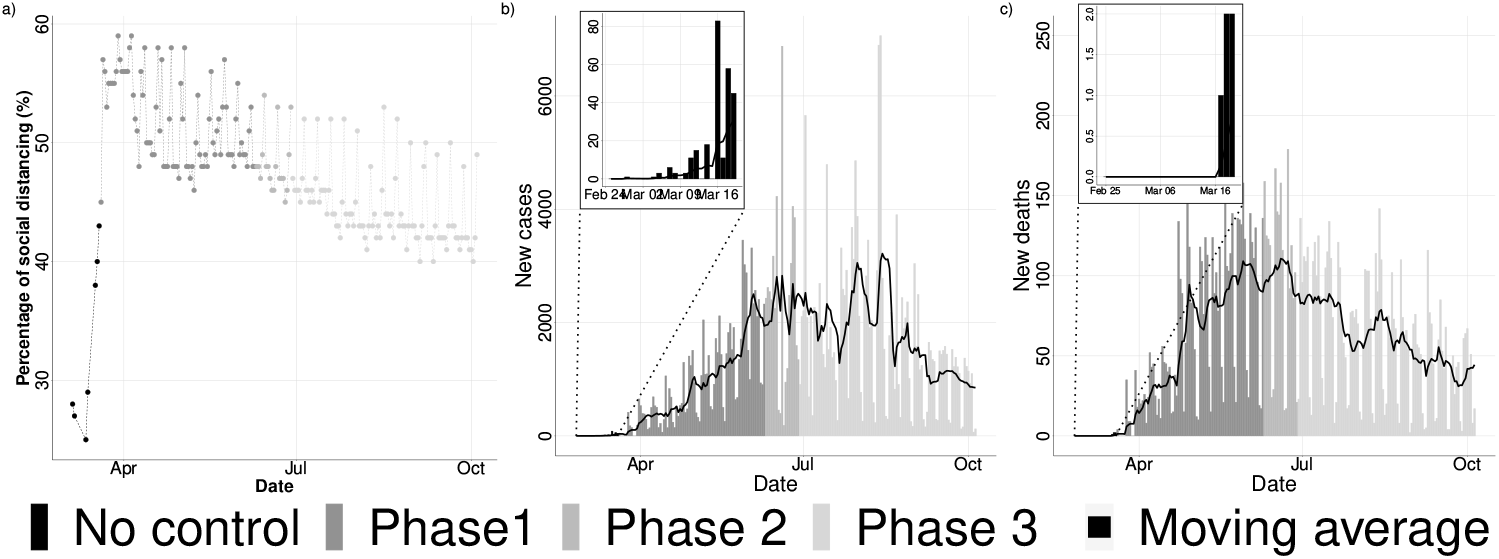
a) Reported percentage of social distancing commitment; b) number of new cases; c) number of new deaths. Data extracted from [19]. The moving average is given by the average of the last seven days of epidemics.

All those features of the dynamics make that a simple SIR model is not able to recover the reported number of cases, or deaths, since its exponential behavior does not achieve a good fitting to the reported numbers. The solution that we proposed here is to fit the transmission rate as a function of time, *β*(*t*), supposing a weekly change for the parameter. The fitting was performed through a genetic algorithm (GA), that is better described in Appendix A, over the reported number of deaths in São Paulo City. Moreover, we excluded the initial days in which the total number of deaths is lower than five.

The result of fitting is showed in Figure 3 that presents the simulated number of deaths compared to the reported number of deaths. While in Figure 4, we show the weekly value of the estimated effective reproductive number, *R*_*t*_, i.e. the average number of secondary cases produced by one single infected individual in the population over the time.

**Figura 3:**
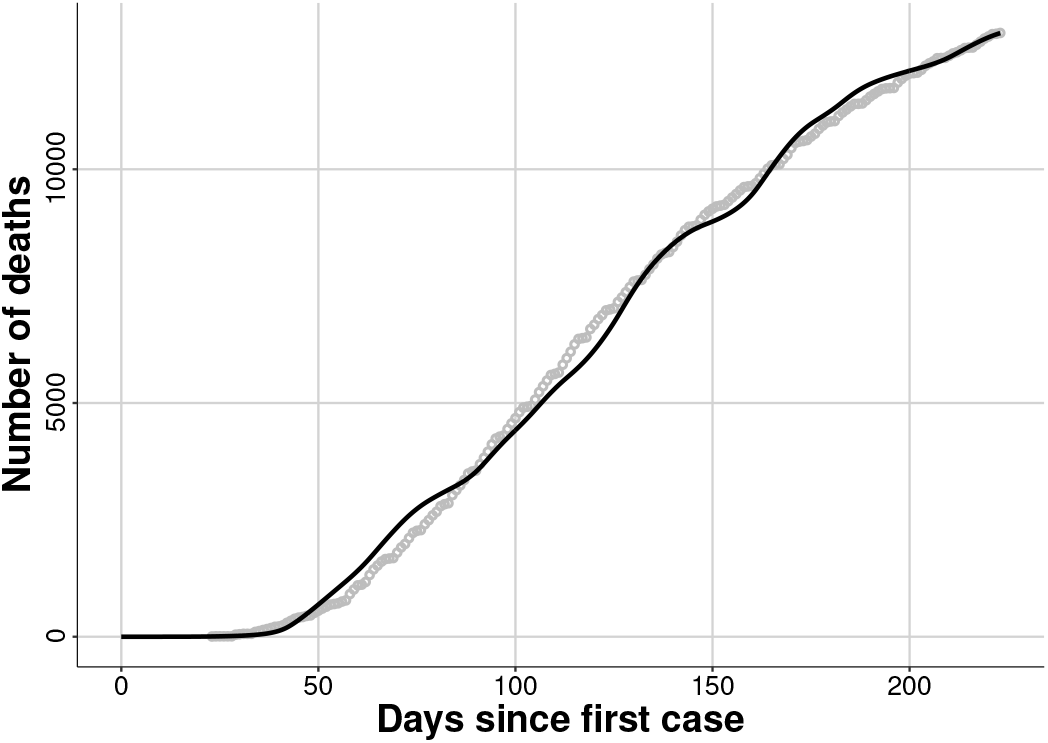
Results of fitting through a genetic algorithm. The comparison between the simulated (continuous line) and the reported cumulative number of deaths (circles).

**Figura 4:**
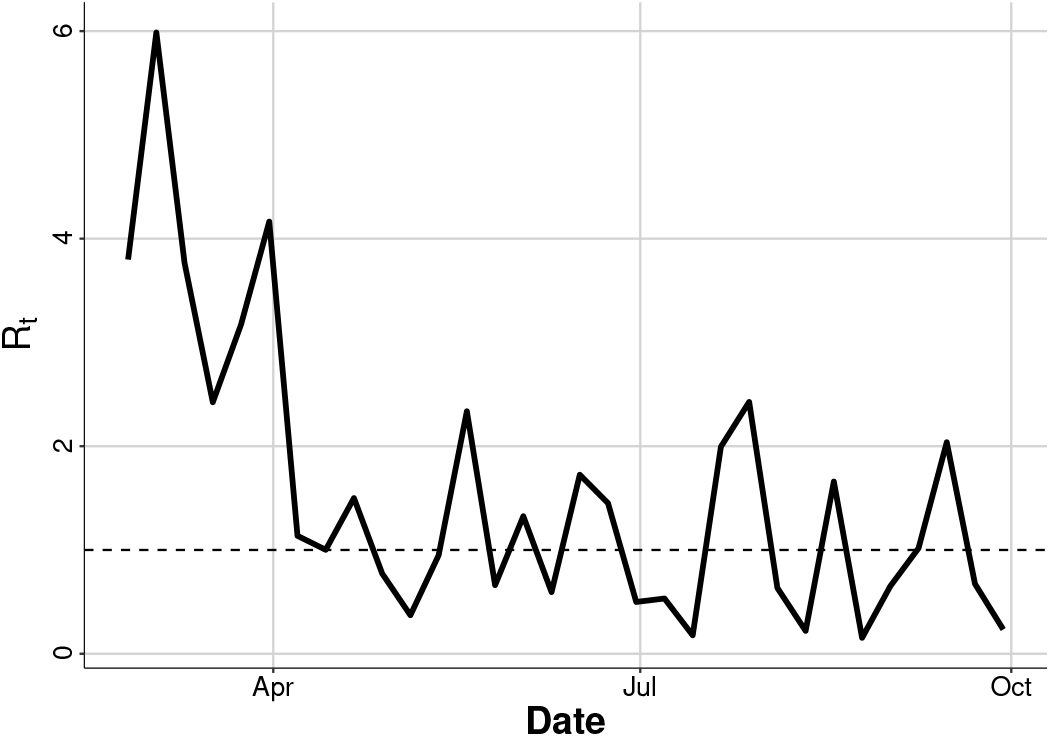
Results of fitting through a genetic algorithm. The estimated effective reproductive number, *R*_*t*_, considering that the herd immunity in population is close to zero. The dashed line emphasizes the value *R*_*t*_ = 1.

### 3.3. Relaxing the Social-Distancing Measures

We simulated two different strategies of the re-opening plan: the first one, Strategy 1, we supposed that every period *p*, São Paulo City changes to the next phase, regardless the epidemic behavior; in the second one, Strategy 2, we supposed that the city evolves to the next phase only if during the previous 15 days, the incidence of infectious individuals decreased everyday (compared to the previous one). We also simulated the scenario in which the system is kept at current phase.

Since June 29^*st*^ (day 125 of the epidemics), São Paulo City is in phase 3. Therefore, we considered, approximately, that the current *β*, called *β*_3_, is given by averaging the last three values of the fitted *β*(*t*), which gives us *β*_3_ = 0.1997 (*R*_*t*_ = 0.98). Then, let’s suppose that, when the city evolves to a next phase, phase 4 and phase 5, *β* is increased by *σβ*_3_, where 0 ≤ *σ* ≤ 1. Which results that, in our simulations,

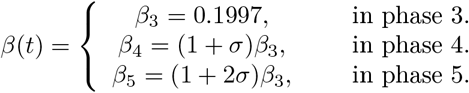

The initial conditions of the simulations are given by the last value of the fitted time series. Then, we simulated the dynamics of the system for the scenarios described above, being the scenario 1 simulated for *p* ∈ (15, 30, 45, 60) days. We present the results of each scenarios in Figures 5 to 7, that show the infectious prevalence, the cumulative number of cases, and the cumulative number of deaths, over the time.

**Figura 5:**
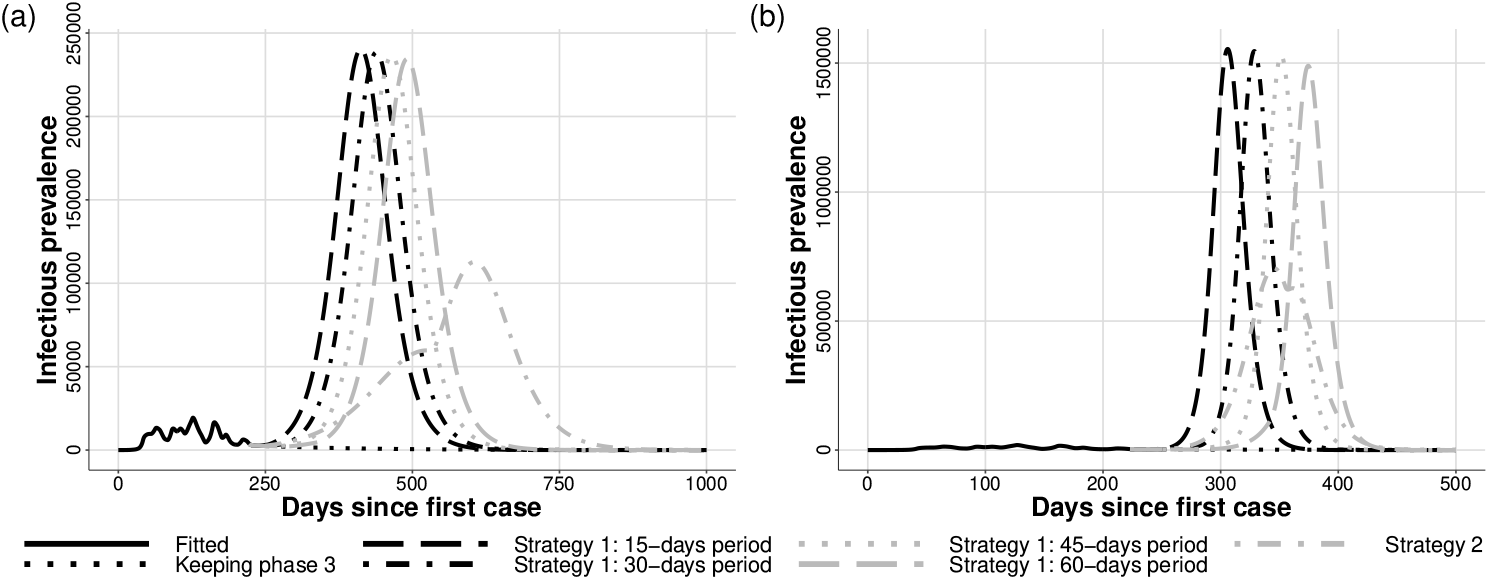
Infectious individuals prevalence. In a) we have *σ* = 0.2 and b) *σ* = 0.8.

**Figura 6:**
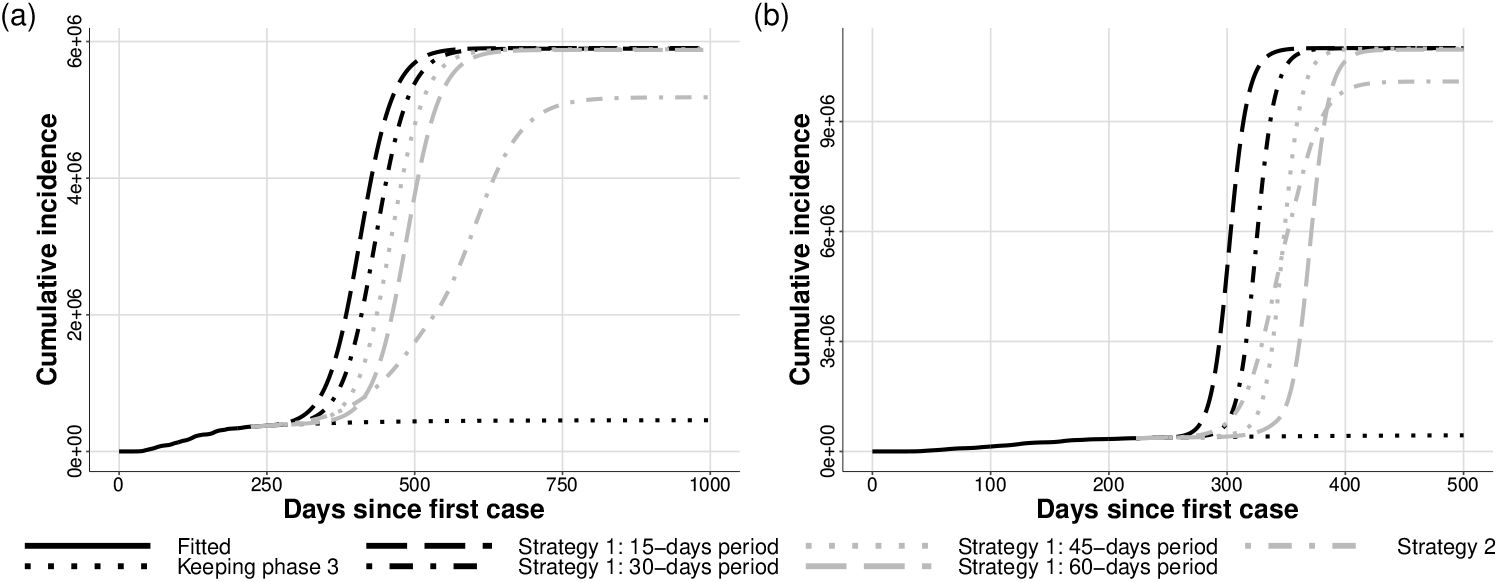
Cumulative number of cases. In a) we have *σ* = 0.2 and b) *σ* = 0.8.

**Figura 7:**
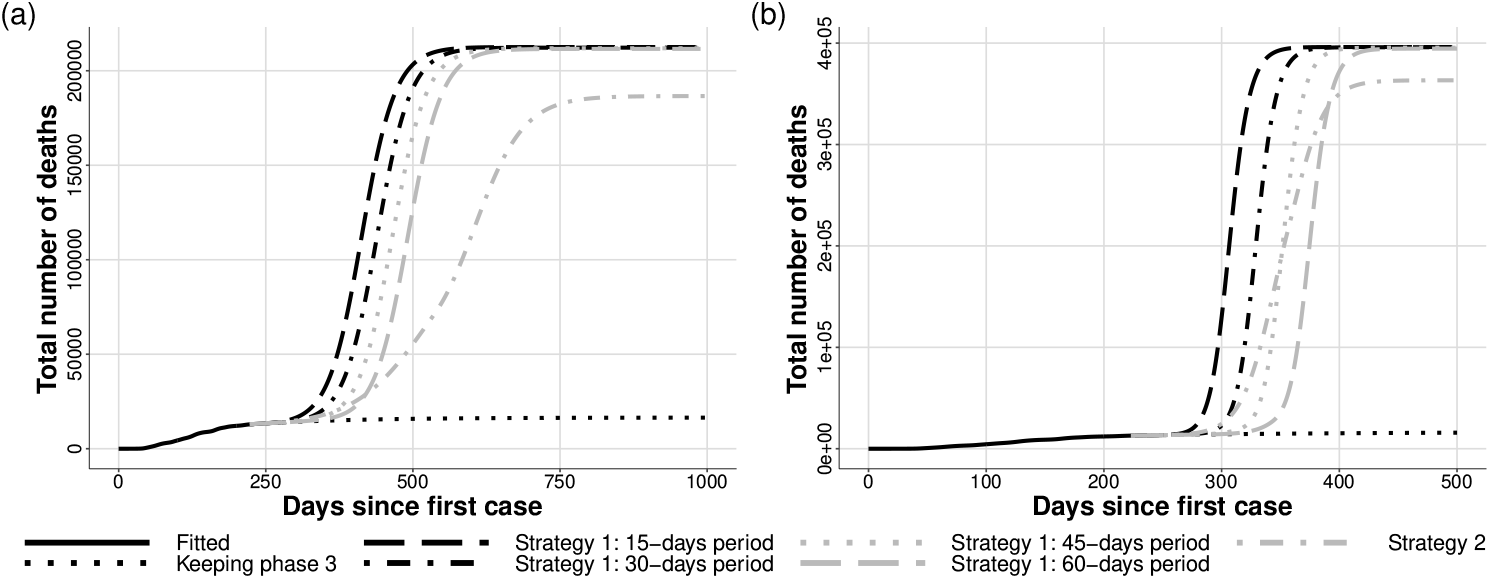
Cumulative number of deaths due to COVID-19. In a) we have *σ* = 0.2 and b) *σ* = 0.8.

We can see that the epidemics peak is not affected by the applied control scenario. Waiting for the incidence of infectious individuals decrease shows to be the strategy with the lowest epidemic peak, as expected. In the same direction, when one looks at the total number of cases and total number of deaths, it is possible to see that all scenarios in Re-Opening Strategy 1 close results, but Strategy 2 was able to decrease those numbers in, around, 12.5%, in both cases (*σ* = 0.2 and *σ* = 0.8.

Since the *R*_*t*_ in Phase 3 is below one, considering the average of weeks from 09-15-2020 to 10-06-2020), but, at the same time, very close to 1, keeping this phase for a large period would slowly decrease the daily number of cases and deaths, and the epidemics would vanish. The dynamics can take more than 250 days to stabilizes.

## 4. Discussion

We presented a simple SEIR model to study the dynamics of the new coronavirus, responsible for the COVID-19. The model considers and simulates the spread of COVID-19, and given that Brazil, and São Paulo State, is still experiencing the first wave of the epidemics, we did not considered a second wave in the model. Moreover, the model considers that a progress to a next phase of the re-opening plan increases the transmission rate, which can happen due to commercial and social activities, given that people may be requested to work and, also, have a decrease of their risk perception.

The model was fitted to data, available officially by the government of São Paulo [19], using a weekly-changing transmission rate, *β*(*t*). The data is shown in Figure 2, while the fitted result and the estimated *R*_*t*_ are in Figures 3 and 4.

Analysing Figures 2 and 4, we see that, after a transient, there is a phase in which the *R*_*t*_ is kept greater than one, corresponding to a phase in which the epidemics strictly grows (between May and June). After that, the *R*_*t*_ goes below one, and remains oscillating around this value, which stabilises the number of daily cases[14]. Given this stabilization, the authorities moved the city to the next phase of the reopening plan, which is reflected, after some weeks (because the intrinsic incubation period), in the number of cases and the *R*_*t*_, according to our estimations. This kind of situation have been experienced in several cities [20]. The city of São Paulo is expected to advance to the SP plan’s green phase until the middle of October 2020 [21].

We simulated two different re-opening strategies, the first one, called Strategy 1, supposing that the city evolves to a less-restrictive phase after a fixed period of 15 to 60 days; and a the second strategy where the city changes the re-opening phase when the incidence of infected individuals decreases for 15 days in a roll. We also simulated the scenario in which SP city keeps phase 3, which is the phase that the city was in the day 224 of epidemics.

Keeping phase 3 would eliminate the epidemic peak that appears when changing phases and decrease the attack rate and the number of deaths; however, it would need several economic activities to stop working in full capacity during more than 250 days. As expected, when increasing the period of phase evolution in scenario 1, the epidemic peak decreases and is delayed; at the end of the epidemics, the attack rate for all simulated situations of Strategy 1 is very similar. When supposing Strategy 2, which is closer to the strategy that the government has been using, the attack rate is controlled to around 12.5% lower.

Despite among reopening scenarios the total attack rate and number of deaths are significantly close to each other, notice that our model does not account to the health system capacity and, therefore, maintain a constant death rate regardless the infectious prevalence, which is directly related to the number of individuals needing hospitalization. However, if the peak of hospitalization needing overcomes the health system’s capacity, the mortality may increase due to the lack of assistance. According to the WHO, 20% of the symptomatic individuals needs healthcare services [22], which, considering that SP City has less than 29,000 available beds [23], may saturate the hospitals of the city if the epidemic peak is large.

This model, as any other one, has limitations. Firstly, it does not account to spatial effects, which may have been addressed as an important characteristic, especially in the low-income communities of SP City [24]. Secondly, we considered that each phase of the re-opening plan increases the transmission rate at the same amount, which may not be true given that some specific activities, such as working-out, may encompass a greater risk of infection. Moreover, we did not account to the health system’s carrying capacity, neither the distinction between asymptomatic and symptomatic infections, which could have a significant impact over the death projection and differ, even more, the tested scenarios.

We emphasize that our goal in this work is not to make predictions, but study the dynamics behavior given some assumptions on the re-opening plan in SP. The main idea is that this study may contribute to authorities justify the current mitigation measures. Despite the results are somehow expected, we do consider that provide simple explanations and analysis for scientific divulgation and decision supporting (or justification) is important. Stochastic, and more predictive, models that take into account more variables, such as asymptomatic, symptomatic individuals, and people who need hospitalization (ward and ICU) merit further investigations.

## Data Availability

The data are available officially by the government of São Paulo

https://www.saopaulo.sp.gov.br/coronavirus/isolamento/

## Author Contributions

Conceptualization: TNV; Methodology: FAR and TNV; Formal analysis and investigation: TNV and FAR; Writing - original draft preparation: FAR; Writing - review and editing: TNV.

## A. Genetic Algorithm

The temporal value of the transmission rate, *β*(*t*) was estimated by fitting the model to the reported number of deaths due to COVID-19. We fixed the parameters as in Table 1 and supposed that *β*(*t*) changes weekly. Finally, we used a genetic algorithm in order to fit the model to the data.

A genetic algorithm is an optimization technique in which an initial population of “chromosomes”is generated and a proportion of them may survive to the next generation or be replaced by a new individual, depending on its fitness score [25].

In our case, each gene of chromosomes in the GA represents the weekly value of *β*(*t*) throughout the simulated time. The fitness score (FS) assigned to each chromosome is given by the inverse of the error between the ODE solution and the number of deaths, as showed in expression A.1.

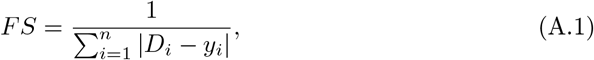

in which *y*_*i*_ is the reported number of deaths on day *i* of the epidemics, and *D*_*i*_ is the solution of the system (2.1) on day *i*, given by a numerical method (fourth-order Runge-Kutta).

The initial population of 1000 chromosomes passed to the GA is created following the given steps:

1. We used the methodology proposed by [26] in order to estimate the effective reproductive number, *R*_*t*_, from data of new cases [14].
2. Using the daily estimation of *R*_*t*_, we calculated the weekly average of *β*(*t*), building a vector ***β*** = [*β*_*i*_].
3. The *i*-th gene of the initial chromosome population is sampled from an exponential distribution based on the estimated value *β*_*i*_.

After that, for each generation of the GA, we performed the following steps:

1. Two chromosomes are randomly selected based on their FS, using a “roulette” system.
2. The two selected chromosomes are randomly broken in one to three points and a crossover happens. It means that two offspring chromosomes are generated.
3. Only the two chromosomes, from the four available ones (two parents and two offspring ones), with the higher fitness are selected to the next generation.
4. This process is performed until the next-generation population has the same population size of the previous one.
5. After that, every gene in population has a probability *p* = 0.005 to suffer a mutation. If it happens, the number is randomly taken from the distribution probability that was used to initialize the population (step 3 of the GA’s initialization).

The GA is run for 600 generations, and for every generation, we drew a boxplot, in Figure 8, for the fitness score over the generations. Since the FS evolves very fast, we plotted the first 30 generations.

**Figura 8:**
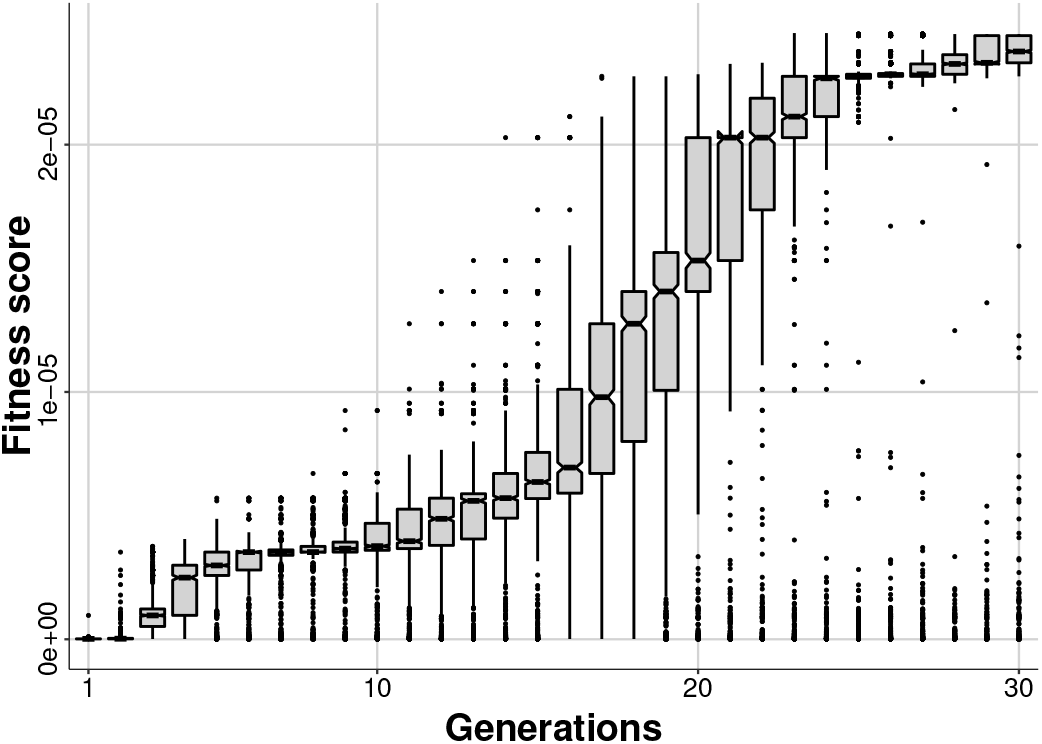
Boxplot showing how the population’s fitness score evolves over the generations of the genetic algorithm.

